# Retrospective mortality and prevalence of SARS-CoV-2 antibodies in greater Omdurman, Sudan: a population–based cross–sectional survey

**DOI:** 10.1101/2021.08.22.21262294

**Authors:** Wendelin Moser, Mohammed Ahmed Hassan Fahal, Elamin Abualas, Shahinaz Bedri, Mahgoub Taj Elsir, Mona Fateh El Rahman Omer Mohamed, Abdelhalim Babiker Mahmoud, Amna Ismail Ibrahim Ahmad, Mohammed A. Adam, Sami Altalib, Ola Adil DafaAllah, Salahaldin Abdallah Hmed, Andrew S. Azman, Iza Ciglenecki, Etienne Gignoux, Alan González, Christine Mwongera, Manuel Albela

## Abstract

**Background:** Even after adjusting for the expected lower severity due to the younger age of the population, relatively low SARS-CoV-2 incidence and mortality rates have been reported throughout Africa. For investigating whether this is truly the case, we conducted a survey to estimate the COVID-19 related mortality and cumulative incidence of SARS-CoV-2 infections in Omdurman the most populated city of the tripartite metropolis Khartoum in Sudan.

**Methods:** A retrospective, cross–sectional, mortality and seroprevalence survey was conducted in Omdurman, Sudan, from March 1, until April 10 2021. A two–stage cluster sampling method was used to investigate the death rate for the pre–pandemic (January 1, 2019–February 29, 2020) and pandemic (March 1, 2020 – day of the survey) period using questionnaires. The seroprevalence survey was performed in a subset of households and all consenting members were tested with a rapid serological test (SD–Biosensor) and a subgroup additionally with ELISA (EUROIMMUN). Fisher’s exact test was used to assess differences between the pre–and pandemic periods and a random effect and Bayesian latent class model to adjust for test performance.

**Findings:** Data from 27315 people (3716 households) for the entire recall period showed a 67% (95% CI 32–110) increase in death rate between the pre–pandemic (0.12 deaths/10000 people/day [95% CI 0.10–0.14]) and pandemic (0.20 [0.16–0.23]) periods. Notably, a 74% (30–133) increase in death was observed among people aged ≥50 years. The adjusted seroprevalence of SARS-CoV-2 was 54.6% (95% CI 51.4–57.8). The seroprevalence was significantly associated with age, increasing up to 80.7% (71.7–89.7) for the oldest age group (≥50 years).

**Interpretation:** Our results showed a significant elevated mortality for the pandemic period with a considerable excess mortality in Omdurman, Sudan. The overall high seroprevalence indicated a different age pattern compared to other countries, with a significant increase by age.

**Funding:** Medécins Sans Frontières

## Introduction

As many key epidemiological and serologic characteristics of severe acute respiratory syndrome coronavirus 2 (SARS-CoV-2) remain unknown, several countries have conducted seroprevalence studies to better understand the extent to which the population has been infected with SARS-CoV-2 at a community level and to monitor its spread over time. Compared with the official number of reported cases, these surveys estimated several–fold higher levels of past infection, reflecting their importance in understanding the full extent of the pandemic across different regions. However, only a few seroprevalence studies have been carried out in Africa, mainly focusing on specific risk groups rather than population–based surveys.^1^ With only limited data, the full extent of COVID-19 and its associated mortality is difficult to estimate and further studies are of great importance for governments and policy makers in order to take decisions on the management of the pandemic.^2^

Contrary to the bleak scenario initially predicted for Africa at the start of the COVID-19 pandemic,^3^ the recorded number of cases and deaths after the first wave remained low compared with other continents, which changed the assumptions towards a scenario where Africa might was the least affected continent. Reasons provided to explain this phenomenon; the younger population structure,^4^ lower prevalence of cardiovascular diseases^5^ and limited capacity for testing.^6^ After the first case was declared in Sudan on March 13, 2020, similar data trends were reported. Only 30404 COVID-19 cases were officially recorded up to April 1, 2021, whereof 72% were registered in Khartoum State alone.^7^ However, between March and July 2020 a survey based on convenience sampling including over 1000 individuals from 22 neighbourhoods of Khartoum city found, that 35% were COVID-19–positive (based on reverse transcriptase–polymerase chain reaction [RT–PCR]), and 18% were positive for anti–SARS-CoV-2 antibodies.^8^ Subsequently, a modelling study for Khartoum State estimated a population seropositivity of 38% and an overall of 16090 undetected COVID-19 deaths up to the end of the first wave by November 2020.^9^

In many low and middle–income settings where access to care and diagnostic tests is limited surveillance systems have not been able to capture the magnitude of the outbreak leading to a discrepancy between the recorded case number and the true extent of SARS-CoV-2 pandemic, as illustrated for Sudan. However, seroprevalence and retrospective mortality studies can fill these data gaps, allowing governments and policy makers to take appropriate decisions on the management of the pandemic. In order to provide complementary information in detail, we conducted two such studies to evaluate the true extend of the SARS-CoV-2 spread and the COVID-19–associated deaths (direct and indirect) in Omdurman, the most populated city of the tripartite metropolis of Khartoum, in order to gain a clearer overall interpretation of the COVID-19 situation.

## Methods

### Study area and design

Sudan’s capital Khartoum is a tripartite metropolis including Khartoum, Bahri and Omdurman with a total of 8 million inhabitants,^9^ located at the confluence of the White and Blue Nile. Omdruman, the largest among the three cities was chosen as study site, which included two surveys: i) a retrospective mortality survey using a two–stage cluster sampling methodology based on random geo–points and ii) a nested SARS-CoV-2 antibody prevalence survey. For the mortality survey the recall period was divided into two periods: the pre–pandemic (January 1, 2019–February 29, 2020) and the pandemic (March 1, 2020–day of the survey) periods.

The primary objective of the survey was to estimate death rate for individuals ≥50 years and the overall seroprevalence of anti–SARS-CoV-2 antibodies in Omdurman. The secondary objectives included, age group– specific (<5, 5–19, 20–34, 35–49, ≥50 years) seroprevalences, risk factors for seropositivity, health seeking behaviour, and access to health care among the people living in greater Omdurman. Additionally, the sensitivity and specificity of the rapid serologic test (RST) was compared to that of the enzyme–linked immunosorbent assay (ELISA).

### Procedures

We used two–stage cluster sampling to select random households. The Ministry of Planning provided a point– file containing each middle–point of polygons representing a residential parcel, which was considered as household for this survey. Proportional to the total number of parcels in the 34 administrative units in greater Omdurman (appendix p 2), 140 points were randomly chosen using the random generator software of ArcGIS 10.5, identifying the first household of a cluster with a total of 30 households. The remaining 29 households were chosen in closest proximity to the first household. All households were included in the mortality survey, whereas the seroprevalence survey included four randomly selected households per cluster among which all family members without any age restriction were invited to participate. Participants were excluded in case of absence after three attempted visits. Additionally, dry blood spots (DBS) were collected from two out of the four households participating in the seroprevalence survey. Previously trained medical doctors forming the survey team carried out the RST and DBS collection.

For the mortality survey, a questionnaire was administered to the head of each household, adapted from the recent WHO recommendations for identifying mortality from COVID-19.^10^ Information gathered included demographics of all household members, details on deceased household members, comorbidities, COVID-19 testing, and health seeking behaviour. For the seroprevalence survey, each participant was asked individually about past symptoms related to COVID-19, ongoing treatment, exposure to a suspect or confirmed COVID-19 case, and other risk factors.

### Sample size calculation

The sample size calculations were performed using the software ENA 2020 (version: Jan/11). The sample size for the mortality survey was based on the mortality of individuals aged ≥50 years (0.73 deaths/10000 people/day),^11^ with a precision of ±0.2, a design effect of 1.2 and a household size of 6 people, resulting in a required 3637 households. For the seroprevalence survey, a SARS-CoV-2 antibody prevalence of 34% in the population was assumed based on the preliminary results from a study in Khartoum.^8^ To allow for age stratification, the sample size was based on the smallest age group (≥50 years), representing approximately 11.5% of the population.^11^ With a precision of ±5%, a 5% type 1 error and 5% inconclusive results, at least 363 individuals per age–group were required.

To assess the diagnostic performance of the RST, a total of 745 samples were required for analysis by ELISA.^12,13^ Specifically, 191 positive and 554 negative samples were needed to confirm the sensitivity (97.0%, precision ±2.5%) and specificity (96.2%, precision: ±1.7%) according to manufacturer.

### Ethics

Ethical approval was obtained from the National Health Review Ethics Committee (No. 3–1–21), Médecins Sans Frontières Ethics Review Board (ID 2089c) and Khartoum State Ministry of Health. Additionally, the three localities of greater Omdurman (Omdurman, Umbedda and Kereri) were informed and authorisation received before seeking authorisation from the administrative units within the localities. Prior to the field data collection, the leader of the “resistance committee” for each block was visited to obtain verbal consent. For the mortality survey, verbal consent was obtained from the head of the household. For the seroprevalence survey, written informed consent was asked from adults and for individuals <18 years, first written informed consent from parents or legal guardians and second, oral assent from the minor itself was obtained.

### Diagnostics

For practical reasons and minimising refusals, the least invasive method with capillary blood collection for rapid serological testing (STANDARD Q COVID-19 IgM/IgG Combo from SD–Biosensor) was selected. All participants either positive for IgM, positive for IgG or positive for IgM and IgG, based on the RST were considered positive for anti–SARS-CoV-2 antibodies. According to the manufacturer, the RST has a sensitivity and specificity of 96.9% (95% CI 91.3–99.4) and 96.2% (93.2–98.2), respectively. The dry blood spot cards (EUROIMMUN, Lübeck, Germany) were transferred to the National Public Health Laboratory (NPHL) in Khartoum for further analysis by ELISA (EUROIMMUN Anti–SARS-CoV-2 ELISA [IgG, S1 domain], Lot: E210118BQ, Lübeck, Germany) following standard operating procedures. Based on the manufacturer the ELISA assay has a sensitivity of 94.4% and a specificity of 99.6% for detecting previous anti-SARS-CoV-2 antibodies.

### Statistical Analysis

The analysis of the data was performed using R (R Core Team, 2020) and Stata V15 (StataCorp. 2017). For the crude death rates (expressed as deaths/10000 people/day) a design effect was assumed to weight the differences among clusters. To compare death rates between the pre–pandemic and pandemic periods the rate ratio was calculated based on a two–sided exact rate ratio test and Fisher’s exact test, was applied to proportions where appropriate. For having the most accurate estimation of the seroprevalence based on the tests used in this survey, two different approaches were defined. First, published performance estimates for the RST were used for a meta-analysis with random effects model (adjustment 1, model description in the appendix p 3). The model provided a corrected estimate of the sensitivity and specificity for adjusting the crude seroprevalence. Considering the lack of a gold standard test for detecting SARS-CoV-2 antibodies, the waning of antibodies and fixed threshold for their detection by the RST,^14,15^ a second adjustment (adjustment 2, model description in the appendix p 4) was done. The survey’s ELISA results were combined with the performance estimation from the previously defined random effects model for both the ELISA and RST and used as inputs for a Bayesian latent-class model (BLCM),^16–18^ resulting in a RST performance estimation used as adjustment. For calculating the beta distributions of the priors for the BLCM, the BetaBuster software^19^ was used. When comparing the results of the RST with ELISA only the positive for IgG or IgG/IgM were considered positive. Risk factors associated with a positive RST were assessed with a logistic regression model. To estimate the excess mortality, SARS-CoV-2 infections and infection fatality rate, the survey results were extrapolated our population estimation - average household size multiplied by the number of households provided by the Ministry of Planning.

## Results

From March 1 until April 10, 2021, a total of 4086 households were visited, thereof 207 (5.1%) refused and 163 (4.0%) were absent after two re–visits (figure 1). In all households participating in the survey, data from 27315 people were recorded. The median age was 22 years (standard deviation [SD] ± 18.4) and an age group as follows: 11.4% (<5 years), 35.1% (5–19 years), 26.5% (20–34 years), 15.0% (35–49 years) and 12.0% (≥50 years).

**Figure 1.**
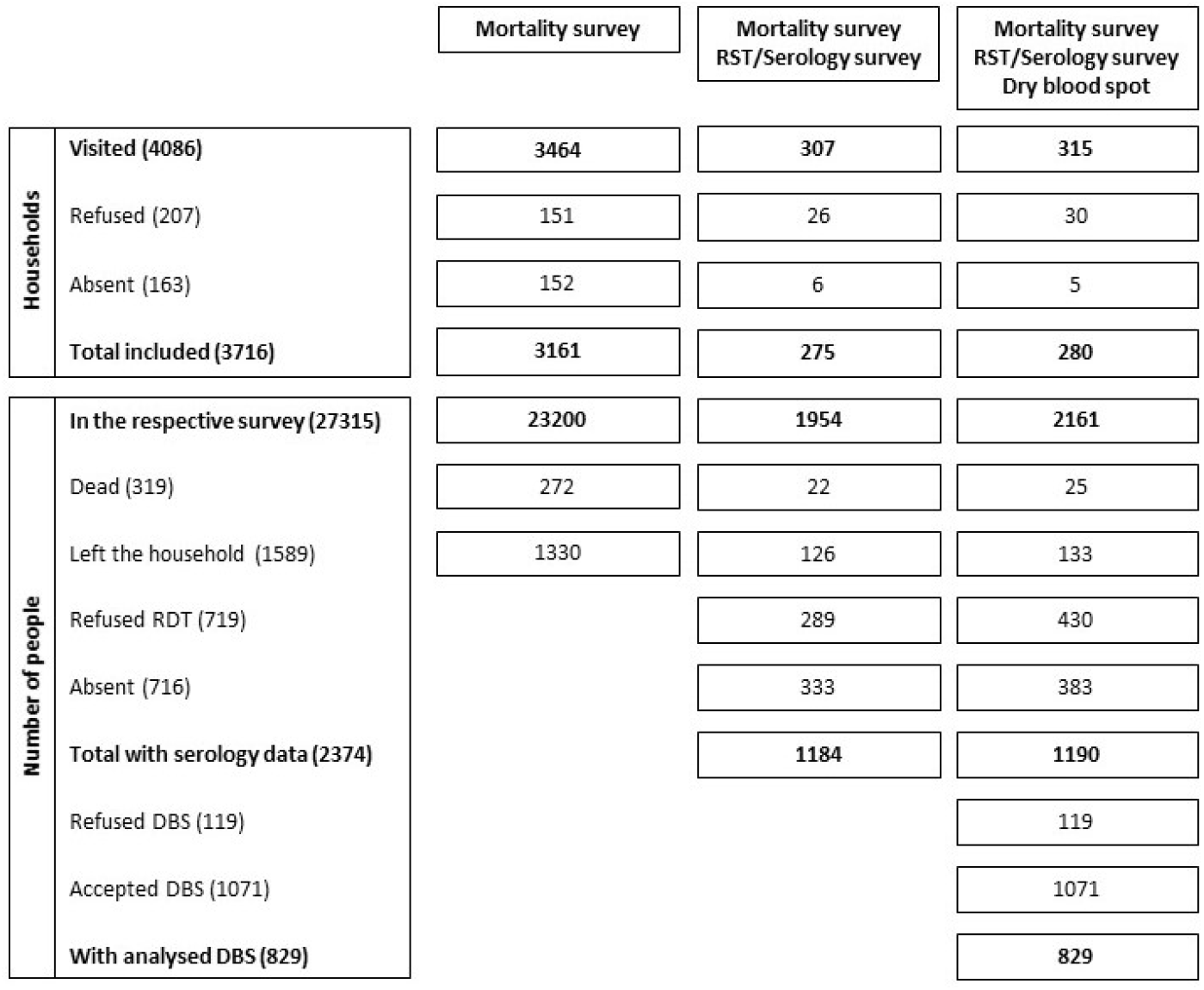
Survey flow. Number of households and people visited; i) mortality survey only, ii) mortality & serology survey with rapid serologic test (RST) only and iii) mortality & serology survey with RST and dry blood spots (DBS).

Among 319 reported deaths, 206 (64.6%) were among individuals aged ≥50 years and 30 (9.4%) deaths were attributed to children below 5 years. The distribution of deaths for the entire recall period (figure 2) showed an overall increasing trend during 2020, in line with the reported countrywide confirmed COVID-19 deaths. This trend was even more pronounced among persons aged ≥50 years for both waves: first during April–May 2020, and then during December 2020–February 2021.

**Figure 2.**
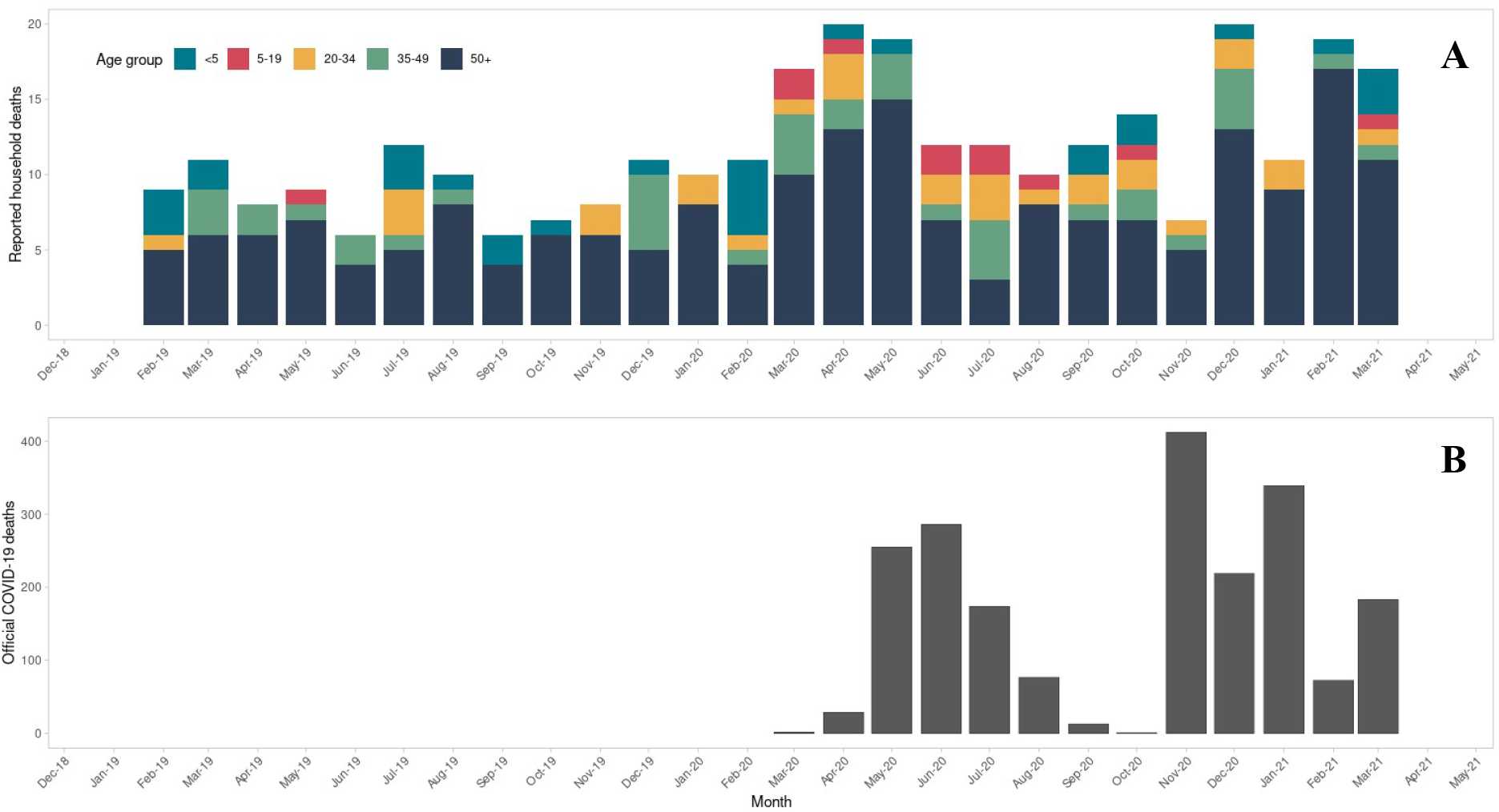
Distribution of the deaths over time. Deaths by age group and month based on the survey (graph A) and deaths per official countrywide registered COVID-19-related deaths (graph B).

The overall death rate for the whole recall period was 0.16 deaths per 10000 people per day (95% confidence interval; 0.13–0.18, table 1). The crude death rate significantly increased by 67% (95% confidence interval 32– 110) from 0.12 (0.10–0.14) for the pre–pandemic to 0.20 (0.16–0.23) for the pandemic period. There was an even more pronounced difference among those aged ≥50 years, with a 74% increase (30–133, p<0.001) between the two periods (0.57 [0.45–0.69] *versus* 0.99 [0.79–1.20]). There was a general upsurge – although not statistically significant – in the death rates for all other age groups, except for children aged <5 years (table 1).

**Table 1.** Death rate for the two periods. Comparison of death rates for the pre-pandemic and pandemic periods across different age groups.

| Age group | Overall |  | Pre-pandemic period |  | Pandemic period |  | Rate ratio |  |
| --- | --- | --- | --- | --- | --- | --- | --- | --- |
|  | n | Death Rate (95% CI) | n | Death Rate (95% CI) | n | Death Rate (95% CI) | Rate ratio (95% CI) | p-value |
| <5 years | 30 | 0.19 (0.10–0.28) | 18 | 0.22 (0.11–0.32) | 12 | 0.17 (0.04–0.30) | 0.77 (0.34–1.70) | 0.613 |
| 5–19 years | 13 | 0.02 (0.01–0.03) | 2 | 0.00 (0.00–0.01) | 11 | 0.03 (0.01–0.05) | – | – |
| 20–34 years | 30 | 0.05 (0.03–0.07) | 10 | 0.04 (0.01–0.06) | 20 | 0.07 (0.04–0.11) | 1.75 (0.78–4.19) | 0.199 |
| 35–49 years | 40 | 0.12 (0.09–0.16) | 16 | 0.09 (0.05–0.14) | 24 | 0.15 (0.09–0.21) | 1.67 (0.85–3.36) | 0.149 |
| $\geq 50$ years | 206 | 0.78 (0.65–0.91) | 80 | 0.57 (0.45–0.69) | 126 | 0.99 (0.79–1.20) | 1.74 (1.30–2.33) | <0.001 |
| Total | 319 | 0.16 (0.13–0.18) | 126 | 0.12 (0.10–0.14) | 139 | 0.20 (0.16–0.23) | 1.67 (1.32–2.10) | <0.001 |

The most common cause of death as reported by the heads of households were non–communicable diseases, followed by trauma/accidents and cancer (appendix p 16), without any differences between the two periods stratified by age groups (appendix p 17). Reported symptoms before death were similar during both periods, apart from more commonly reported extreme fatigue and muscle pain during the pandemic period (appendix p 18). No differences between the two periods for comorbidities rates (appendix p 19), access to health facilities and place of death (appendix p 20) were seen.

3808 individuals were included in the seroprevalence part of the survey (figure 1). Among those, 2374 agreed (62.3%), 719 (18.9%) refused within the household, and 716 (18.8%) were absent during all re–visits; the latter comprised mainly males of working age (appendix p 21). 34.3% (95% confidence interval 32.4–36.2, table 2) of all participants were tested positive for SARS-CoV-2 antibody (IgM and/or IgG). After adjusting the seroprevalence the overall estimates (table 2) increased to 44.0% (41.5-46.6, adjustment 1) and 54.6 (51.4-57.8, adjustment 2). Lowest crude seroprevalence was observed in the youngest age group (<5 years) with 18.7% (14.7–23.5) and significantly increased by age (Odds Ratio 1.01 [1.01–1.02], appendix p 22). The highest crude seroprevalence (p<0.01 compared to all age groups) was observed for the oldest age group (≥50 years) at 50.2% (44.7–55.6). After adjustments, the significant highest estimates remained for people ≥50 years with 65.0% (57.8-72.2, adjustment 1) and 80.7 %(71.7-89.7, adjustment 2). Among those who tested positive, the majority presented IgG antibodies (84.9%, appendix p 23), whereas 10.6% tested positive for both antibodies and 4.5% for IgM only.

**Table 2.** SARS-CoV-2 seroprevalence. Summary of the SARS-CoV-2 antibody seroprevalence according to rapid serologic test results (RST), adjustment 1 (based on RST literature) and adjustment 2 (based on Bayesian latent class model). P–values indicate the difference in relative risk between the oldest age group (≥50 years) as reference and the other age groups.

| Age Group | Rapid Serological Test Result |  |  | Adjustment 1 (based on RST literature) |  |  | Adjustment 2 (based on the Bayesian latent class model) |  |  |
| --- | --- | --- | --- | --- | --- | --- | --- | --- | --- |
|  | Seroprevalence (95% CI) | Relative Risk (95% CI) | P-Value | Seroprevalence (95% CI) | Relative Risk (95% CI) | P-Value | Seroprevalence (95% CI) | Relative Risk (95% CI) | P-Value |
| <5 years (n=299) | 18.7 (14.7-23.5) | 0.4 (0.3-0.5) | <0.001 | 23.5 (18.1-29.8) | 0.3 (0.2-0.4) | <0.001 | 29.0 (22.4-36.9) | 0.3 (0.3-0.4) | <0.001 |
| 5–19 years (n=786) | 30.6 (27.5-33.9) | 0.6 (0.5-0.7) | <0.001 | 39.1 (35.0-43.5) | 0.6 (0.5-0.7) | <0.001 | 48.5 (43.3-53.9) | 0.6 (0.5-0.6) | <0.001 |
| 20–34 years (n=629) | 35.5 (31.8-39.3) | 0.7 (0.6-0.8) | <0.001 | 45.6 (40.8-50.6) | 0.7 (0.6-0.8) | <0.001 | 56.5 (50.5-62.8) | 0.7 (0.6-0.7) | <0.001 |
| 35–49 years (n=342) | 39.5 (34.4-44.7) | 0.8 (0.7-0.9) | 0.006 | 50.9 (44.2-57.9) | 0.8 (0.7-0.9) | 0.002 | 63.1 (54.8-71.8) | 0.8 (0.7-0.9) | <0.001 |
| $\geq 50$ years (n=319) | 50.2 (44.7-55.6) | Reference | | 65.0 (57.8-72.2) | Reference | | 80.7 (71.7-89.7) | Reference | |
| Overall (n=2,375) | 34.3 (32.4-36.2) |  |  | 44.0 (41.5-46.6) |  |  | 54.6 (51.4-57.8) |  |  |

825 samples were tested with the ELISA, of which 244 (29.6%) and 322 (39.0%) where positive for SARS-CoV-2 IgG by the RST and ELISA respectively (appendix p 24). Among 198 cases with discordant results considering ELISA as standard, 60 (30.0%) were false positives and 138 (70.0%) false negatives according to the RST. Based on adjustment 2 including the BLCM, the RST had a sensitivity and a specificity of 64.9% (95% confidence interval 53.9–81.0) and 95.8% (91.0–99.4), respectively (appendix p 25).

Other than age, living with person who was seropositive led to a 1.68 (Odds Ration [OR] 95% confidence interval 1.35–2.08, p<0.001 appendix p 22) fold increase in the odds of being seropositive. Among all 555 included households, 364 (65.6%) had at least one positive household member and 203 household (36.6%) at least two. Sex was not a significant predictor for seroprevalence (p=0.127, appendix p 22).

Using the average household size of 6.9 persons (95% CI, 6.8-7.0) based on the collected data, an estimated 3040604 (95% CI, 2991847–3089359) people live in Omdurman (table 3). We estimate excess number of deaths to 7113 deaths (5015–9505) for the overall population and 5125 deaths (4165–6226) among people aged ≥50 years. The estimated number of people with a past SARS-CoV-2 infection in Omdurman is 1040765 (95% confidence interval 920674-1166662) and 1660170 (1458225-1863936) based on the crude and second adjustment of the seroprevalence, respectively.

**Table 3.**
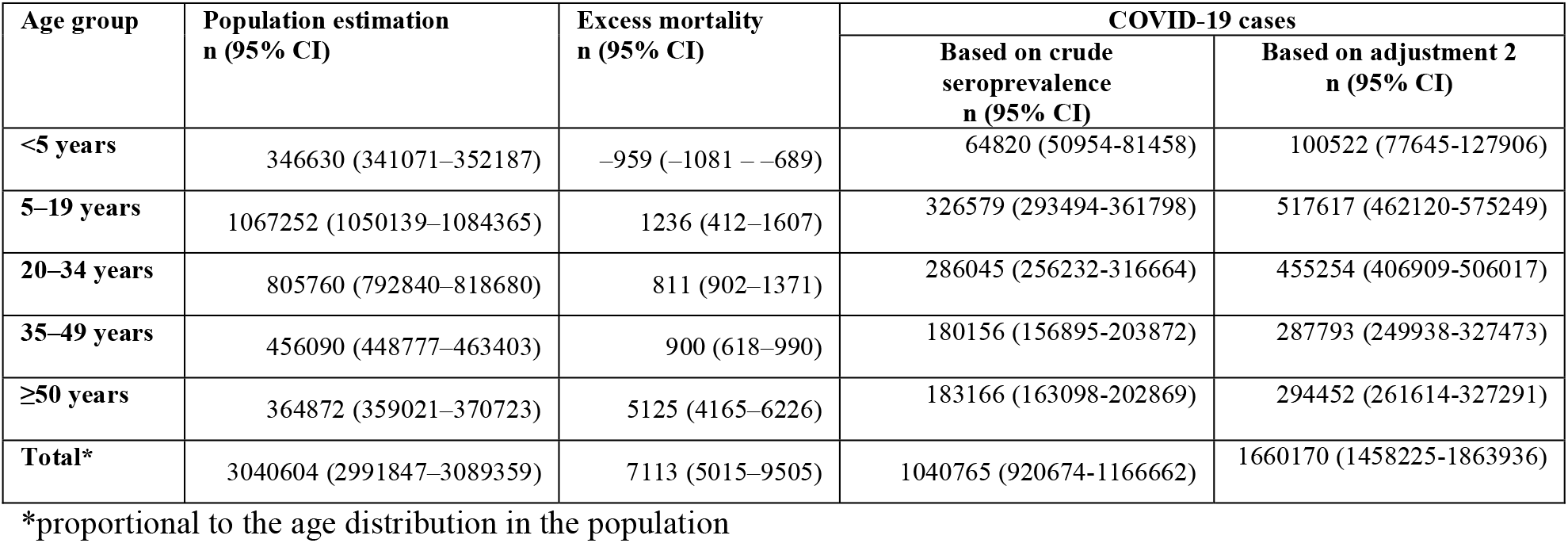
Excess mortality and estimated number of COVID-19 cases by age group.

| Age group | Population estimation<br>n (95% CI) | Excess mortality<br>n (95% CI) | COVID-19 cases |  |
| --- | --- | --- | --- | --- |
|  |  |  | Based on crude<br>seroprevalence<br>n (95% CI) | Based on adjustment 2<br>n (95% CI) |
| <5 years | 346630 (341071–352187) | –959 (–1081 – –689) | 64820 (50954–81458) | 100522 (77645–127906) |
| 5–19 years | 1067252 (1050139–1084365) | 1236 (412–1607) | 326579 (293494–361798) | 517617 (462120–575249) |
| 20–34 years | 805760 (792840–818680) | 811 (902–1371) | 286045 (256232–316664) | 455254 (406909–506017) |
| 35–49 years | 456090 (448777–463403) | 900 (618–990) | 180156 (156895–203872) | 287793 (249938–327473) |
| $\geq 50$ years | 364872 (359021–370723) | 5125 (4165–6226) | 183166 (163098–202869) | 294452 (261614–327291) |
| **Total\*** | 3040604 (2991847–3089359) | 7113 (5015–9505) | 1040765 (920674–1166662) | 1660170 (1458225–1863936) |

\*proportional to the age distribution in the population

## Discussion

This survey is among the very first, detailed studies investigating COVID-19 mortality alongside anti–SARS-CoV-2 antibody prevalence in Africa. In Omdurman, we found an overall increased mortality for the pandemic period and high levels of SARS-CoV-2 infections particularly in the ≥50 age group.

Our findings indicate a significant increase of 67% in deaths for the overall population of Omdurman during the pandemic period, with the highest increase among the older population aged ≥50 years (74%). The observed mortality trends for Omdurman followed similar pattern as the official recorded countrywide COVID-19 deaths; Peak months coinciding with the first (April–May) and second (December 2020–March 2021) waves in Sudan.

These peaks were corroborated by an increase in burials during the first wave, anecdotal reports on community deaths during both waves (on social media), an estimated 16090 COVID-19 related deaths for Khartoum State until end of November 2021,^9^ and overwhelmed hospitals lacking beds and ventilators during of these peak periods.^7^ The overall death rate for the pre–pandemic period (0.12 deaths/10000 people/day) in Omdurman was lower than the respective country-wide estimates of 0.17 from the United Nations.^11^ Likewise, the death rate for people ≥50 years in Omdurman (0.57) was lower than recorded for the whole country. Similar data for Khartoum State is not available, but we would expect it to be below the countrywide estimate due to higher access to health care in urban settings. These discrepancies between Sudan and Omdurman figures could also be due to the potential limitation of over two years long recall period for mortality estimates, possibly introducing a bias for deaths occurring at the beginning of the recall period. Surveyors were trained to be aware of this factor to mitigate potential recall bias.

The crude seroprevalence shows how widespread the SARS-Cov-2 infection was, affecting all age groups, especially individuals aged 50 years and older. However, the estimates based on the RST might have underestimated the seroprevalence due to several limitations. Firstly, as our survey was undertaken one year after the first SARS-Cov-2 case was detected in Sudan, one can expect a varying degree of antibody decay over time.^20,21^ Second, when antibodies remain present in the blood, their detection is limited by the performance of the RST.^22^ To overcome these limitations, two adjustments of the seroprevalence based on the RST were done. After the first adjustment an almost 10% increase (34.3 to 44.0%) in the overall seroprevalence was observed. An even higher increase of 20% in the overall seroprevalence (34.2% to 54.6%), resulted from the second adjustment. This highlights the importance of considering the limitations of the tests used in seroprevalence surveys and the necessity to consider existing performance data for the tests used. Despite the second adjustment included more information than the first one and should be considered as most accurate estimation, it is important to also highlight its limitations. The priors used for BLCM were based on performance data from various settings and populations around the world, which may differ from Omdurman. However, given the mix of contexts included in these studies, we consider that the effect should be minimal. The real seroprevalence in Omdurman may lay between the two adjustments which goes in line with a modelling study which estimated a 38% seroprevalence by the end of the first wave (November 2020) in Khartoum.^9^

Compared to global observed trends of higher seroprevalence in the working age population (18-64 years),^23^ we found a different SARS-CoV-2 infection pattern with a gradual increase by age (appendix p 26) up to highest adjusted estimates (adjustment 2) among individuals aged ≥50 years (80.7%). Studies from the neighbouring country’s capital Juba in South Sudan^24^ and from Cameroon^25^ showed a similar, but less pronounced increase by age. Closest to our findings, an overall seroprevalence of 58.5% was found in Mali, which was equally associated with age.^26^ This gradual increase in seroprevalence in Sudan may be explained by various cultural aspects, such as elderly persons’ role in traditional and religious activities (e.g. weddings) which obliges them to be present at social gathering, thereby increasing their number of contacts. Although the concept of shielding the vulnerable elders in the context of COVID-19 in Sudan is acceptable according to cultural norms, its implementation outside of the household is challenging due to social stigma and potential loss of income,^27^ an aspect which may also be reflected in our data.

Various factors have been proposed to explain the overall low numbers of COVID-19 cases and deaths reported from African countries, supporting the assumption that Africa might be less affected by the pandemic.^6^ However, our data contradicts this notion, with the overall seroprevalence detected for Omdurman being amongst the highest compared to all compiled studies listed on the SeroTracker from Arora et al.^1^ Furthermore, considering the high seroprevalences from South Sudan, Cameroon and Mali,^24–26^ the officially reported case numbers in Africa seem to be highly underreported. For example, a study in Zambia showed a 100 times higher number cases compared to the officially reported cases.^28^ Likewise for our study, based on the crude and adjusted (adjustment 2) seroprevalence, the number of people with a past SARS-CoV-2 infection was 1040765 and 1660170 in Omdurman, respectively, compared to the officially reported 5672 COVID-19 cases (data from the Khartoum State Ministry of Health) up to April 10, 2021. Similar trends were observed for the estimated excess of 7113 deaths for the pandemic period, compared to 287 deaths (data from the Khartoum Ministry of Health) officially reported for Omdurman. Our survey together with the few other studies in African countries indicate, that the impact of COVID-19 in the continent might be underestimated and highlight the need for more studies investigating the mortality and seroprevalence to understand the real burden of COVID-19.

In conclusion, our population-based cross-sectional survey in Omdurman, Sudan, demonstrated significantly higher death rate compared to pre-pandemic period particularly affecting individuals aged 50 years and above. We also found elevated seropositivity in Omdurman, with the oldest population range being the most affected. Our results suggest that the African city of Omdurman was severely impacted by the COVID-19 pandemic.

## Data Availability

The datasets used and/or analyses underlying the findings of this survey are available on request, in accordance with the Ministry of Health of Sudan and the legal framework set forth by Médecins Sans Frontiéres (MSF) data sharing policy. MSF is committed sharing and disseminate health data from its programs and research in an open, timely, and transparent manner in order to promote health benefits for populations while respecting ethical and legal obligations towards patients, research participants, and their communities. The MSF data sharing policy ensures that data will be available upon request to interested researchers while addressing all security, legal, and ethical concerns.

## Contributors

All authors contributed to the interpretation of data, and the review and approval of the final manuscript.

## Data sharing

The datasets used and/or analyses underlying the findings of this survey are available on request, in accordance with the Ministry of Health of Sudan and the legal framework set forth by Médecins Sans Frontières (MSF) data sharing policy.^29^ MSF is committed sharing and disseminate health data from its programs and research in an open, timely, and transparent manner in order to promote health benefits for populations while respecting ethical and legal obligations towards patients, research participants, and their communities. The MSF data sharing policy ensures that data will be available upon request to interested researchers while addressing all security, legal, and ethical concerns.

## Acknowledgments

We are thankful to all people participating in the survey, the survey surveyors for their hard work in the field, the laboratory technicians from the National Public Health Laboratory for the ELISA analysis, the team of the Innovation, Development and Research Directorate of the State Ministry of Health for their support, and Tania Kapoor for editorial support (funded by MSF). We would like to acknowledge the National Public Health Laboratory for the donation of the 5000 RST for the survey with kind permission from the Africa Centre for Disease Control.

