## Supplementary Appendix for "Retrospective mortality and prevalence of SARS-CoV-2 antibodies in greater Omdurman, Sudan: a population–based cross–sectional survey"

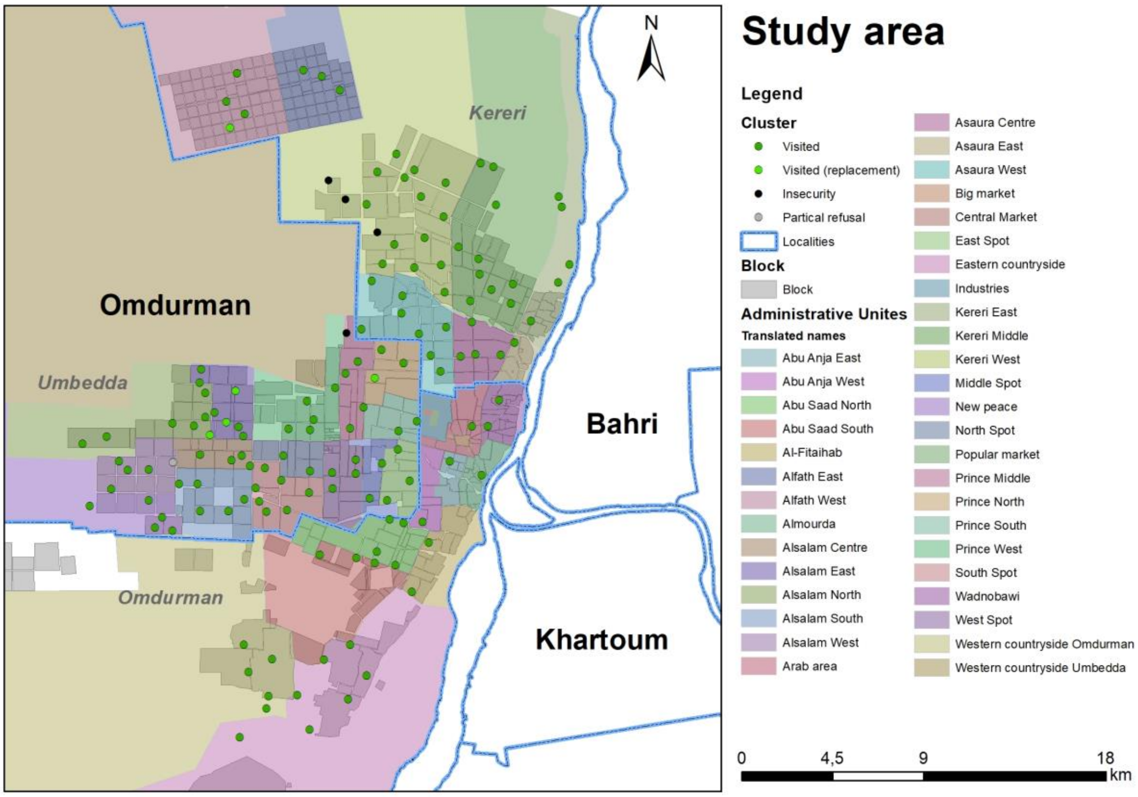


**Figure S1. Distribution of the clusters in the 34 administrative units in greater Omdurman, Sudan**

### Adjustment 1 – Meta-analysis with random effects model

For the adjustment 1, the sensitivity and specificity of the tests was estimated using a univariate random effects model following the methods described by Shim SR *et al.*[1] The idea behind this model is to account for the existing performance data for the tests to improve the estimation of the results. For the model a meta-analysis of the existing published performance data was done for each of the test (rapid serological test [RST] and ELISA) using the "Metaprop" function from the R package Meta. For the e sensitivity and specificity estimation of the RST we gather data from 7 studies. Overall, 6 studies derived from the FIND (the global alliance for diagnostics) database[2] and data were filtered by assay name (STANDARDTM Q COVID-19 IgM/IgG Combo Test), target (IgG/IgM) and all periods. The last study was from the US Food and Drug Administration (FDA) independent evaluations of COVID-19 serological tests[3]. For the ELISA analysis four studies were used: two independent,[4,6] one from the US FDA[5] and the fourth one from the FIND database[7]. The resulting sensitivity and specificity for the RST were 76.6% (95% CI 67.8-83.6) and 99.0% (98.3-99.4) and for ELISA 76.9% (61.1-87.6) and 99.1% (97.7-99.7), respectively. To estimate the adjusted seroprevalence we used the "epi.prev" function from the R package EpiR with the estimated specificity and sensitivity of the RST resulted from the meta-analysis.

### Adjustment 2 – Bayesian latent class model

The adjustment 2 was used to account for the lack of a goldstandard for the detection of SARS-CoV-2 antibodies and the use of imperfect tests. For this purposes we used a Bayesian Latent Class Model (BLCM) based on the Hui-Walter paradigm which has been widely documented[8–10] as a valuable tool to estimate the performance of diagnostic tests in scenarios similar to the ones experienced in this survey. To develop this model we used the "runjags" R package[11] and the data of the participants with a ELISA and RST tests result. The data was divided into five groups according to the age . The BetaBuster[12] software was used to calculate the beta distributions of the sensitivity and specificity resulted from the previous meta-analysis. This information resulted in the following prior beta functions and used as parameters for the BLCM: RST sensitivity dbeta(61.22,19.40), specificity dbeta(965.86, 10.74); ELISA sensitivity dbeta(21.77, 7.24), specificity dbeta(332.21, 4.01). Based on this model we estimated a RST sensitivity and specificity of 61.9% (56.8-66.9) and 98.9% (98.2-99.5). To estimate the adjusted seroprevalence we used the "epi.prev" function with the dataset of all the participants in the serology part of the survey and the estimated specificity and sensitivity of the RST from the BLCM.

**Adjustment 1 – R Code**

RST

```
## Studies 1-6 <- https://www.finddx.org/sarscov2-eval-antibody/
## Study 7 <- https://open.fda.gov/apis/device/covid19serology/
https://www.accessdata.fda.gov/cdrh_docs/presentations/maf/maf3274-a001.pdf

S1sens <- 0.667
S1spec <- 1
S1pos <- 96
S1neg <- 102
TP1 <- round(S1sens*S1pos)
TN1 <- round(S1spec*S1neg)
FN1 <- S1pos-TP1
FP1 <- S1neg-TN1
S2sens <- 0.719
S2spec <- 0.991
S2pos <- 166
S2neg <- 196
TP2 <- round(S2sens*S2pos)
TN2 <- round(S2spec*S2neg)
FN2 <- S2pos-TP2
FP2 <- S2neg-TN2
S3sens <- 0.577
S3spec <- 0.976
S3pos <- 317
S3neg <- 125
TP3 <- round(S3sens*S3pos)
TN3 <- round(S3spec*S3neg)
FN3 <- S3pos-TP3
FP3 <- S3neg-TN3
S4sens <- 0.820
S4spec <- 0.990
S4pos <- 579
S4neg <- 423
TP4 <- round(S4sens*S4pos)
TN4 <- round(S4spec*S4neg)
FN4 <- S4pos-TP4
FP4 <- S4neg-TN4
S5sens <- 0.807
S5spec <- 0.996
S5pos <- 262
S5neg <- 298
TP5 <- round(S5sens*S5pos)
TN5 <- round(S5spec*S5neg)
FN5 <- S5pos-TP5
FP5 <- S5neg-TN5
S6sens <- 0.897
S6spec <- 0.984
S6pos <- 483
S6neg <- 321
TP6 <- round(S6sens*S6pos)
TN6 <- round(S6spec*S6neg)
FN6 <- S6pos-TP6
FP6 <- S6neg-TN6
S7sens <- 0.763
S7spec <- 0.988
S7pos <- 30
S7neg <- 80
TP7 <- round(S7sens*S7pos)
TN7 <- round(S7spec*S7neg)
```

```
FN7 <- S7pos-TP7
FP7 <- S7neg-TN7

AuditC5 <- data.frame(TP = c(TP1, TP2, TP3, TP4, TP5, TP6, TP7),
                      FN = c(FN1, FN2, FN3, FN4, FN5, FN6, FN7),
                      FP = c(FP1, FP2, FP3, FP4, FP5, FP6, FP7),
                      TN = c(TN1, TN2, TN3, TN4, TN5, TN6, TN7))
AuditC5$names <- c("Study 1", "Study 2", "Study 3","Study 4", "Study 5", "Study 6", "Study 7")

sens_logit_rdt <- metaprop(AuditC5$TP, AuditC5$TP + AuditC5$FN, comb.fixed=F, comb.random=T, sm="PLOGIT", method = "GLMM", method.ci="CP", studlab=AuditC5$names)

spec_logit_rdt <- metaprop(AuditC5$TN, AuditC5$TN + AuditC5$FP, comb.fixed=FALSE, comb.random=TRUE, sm="PLOGIT", method.ci="CP", studlab=AuditC5$names)
```

<u>Outputs</u>

```
        proportion         95%-CI
Study 1      0.667 [0.563; 0.760]
Study 2      0.717 [0.642; 0.784]
Study 3      0.577 [0.521; 0.632]
Study 4      0.820 [0.787; 0.851]
Study 5      0.805 [0.752; 0.851]
Study 6      0.896 [0.866; 0.922]
Study 7      0.767 [0.577; 0.901]

Number of studies combined: k = 7

                     proportion         95%-CI
Random effects model      0.766 [0.678; 0.836]

Quantifying heterogeneity:
 tau^2 = 0.3118; tau = 0.5584; I^2 = 95.0% [91.9%; 96.9%]; H = 4.48 [3.52; 5.70]

Test of heterogeneity:
      Q d.f.  p-value             Test
 120.58    6 < 0.0001        Wald-type
 126.95    6 < 0.0001 Likelihood-Ratio
```

**Figure S2: Output of the metaprop function for the estimation of the sensitivity for the RST**

```
        proportion         95%-CI
Study 1      1.000 [0.964; 1.000]
Study 2      0.990 [0.964; 0.999]
Study 3      0.976 [0.931; 0.995]
Study 4      0.991 [0.976; 0.997]
Study 5      0.997 [0.981; 1.000]
Study 6      0.984 [0.964; 0.995]
Study 7      0.988 [0.932; 1.000]

Number of studies combined: k = 7

                     proportion         95%-CI
Random effects model      0.990 [0.983; 0.994]

Quantifying heterogeneity:
 tau^2 = 0; tau = 0; I^2 = 0.0% [0.0%; 52.4%]; H = 1.00 [1.00; 1.45]

Test of heterogeneity:
    Q d.f. p-value             Test
 3.68    6  0.7198        Wald-type
 6.52    6  0.3675 Likelihood-Ratio
```

**Figure S3: Output of the metaprop function for the estimation of the specificity of the RST**

```
##Estimation of the adjusted seroprevalence with a given sensitivity and specificity of a test
adj_serop_overall <- epi.prev(nrow(sero[test_result_cat=="Positive", ]),
                    nrow(sero[!is.na(test_result_cat) & outcome == "done", ]),
                    se = 0.766, sp = 0.990)
adj_serop_o50 <- epi.prev(nrow(sero[age_group == "[50,Inf)" & test_result_cat=="Positive", ]),
                    nrow(sero[age_group == "[50,Inf)" & !is.na(test_result_cat) & outcome == "done", ]),
                    se = 0.766, sp = 0.990)
adj_serop_35_50 <- epi.prev(nrow(sero[age_group == "[35,50)" & test_result_cat=="Positive", ]),
                    nrow(sero[age_group == "[35,50)" & !is.na(test_result_cat) & outcome == "done", ]),
                    se = 0.766, sp = 0.990)
adj_serop_20_35 <- epi.prev(nrow(sero[age_group == "[20,35)" & test_result_cat=="Positive", ]),
                    nrow(sero[age_group == "[20,35)" & !is.na(test_result_cat) & outcome == "done", ]),
                    se = 0.766, sp = 0.990)
adj_serop_5_20 <- epi.prev(nrow(sero[age_group == "[5,20)" & test_result_cat=="Positive", ]),
                    nrow(sero[age_group == "[5,20)" & !is.na(test_result_cat) & outcome == "done", ]),
                    se = 0.766, sp = 0.990)
adj_serop_0_5 <- epi.prev(nrow(sero[age_group == "[0,5)" & test_result_cat=="Positive", ]),
                    nrow(sero[age_group == "[0,5)" & !is.na(test_result_cat) & outcome == "done", ]),
                    se = 0.766, sp = 0.990)
```

### ELISA

```
## Calculating the sens and spec of the ELISA test through a meta-analysis
## Study 1 <- Kr?ttgen A, Cornelissen CG, Dreher M, Hornef M, Im?hl M, Kleines M. Comparison of four new commercial serologic assays for determination of SARS-CoV-2 IgG. J Clin Virol. 2020;128:104394. doi:10.1016/j.jcv.2020.104394
## Study 2 <- https://open.fda.gov/apis/device/covid19serology/ https://www.accessdata.fda.gov/cdrh_docs/presentations/maf/maf3246-a001.pdf
## Study 3 <- Kohmer N, Westhaus S, R?hl C, Ciesek S, Rabenau HF. Brief clinical evaluation of six high-throughput SARS-CoV-2 IgG antibody assays. J Clin Virol. 2020;129:104480. doi:10.1016/j.jcv.2020.104480
## Study 4 <- https://www.finddx.org/sarscov2-eval-antibody/

S1sens_el <- 0.864
S1spec_el <- 0.962
S1pos_el <- 22
S1neg_el <- 53
TP1 <- round(S1sens_el*S1pos_el)
TN1 <- round(S1spec_el*S1neg_el)
FN1 <- S1pos_el-TP1
FP1 <- S1neg_el-TN1
S2sens_el <- 0.900
S2spec_el <- 1
S2pos_el <- 30
S2neg_el <- 80
TP2 <- round(S2sens_el*S2pos_el)
TN2 <- round(S2spec_el*S2neg_el)
FN2 <- S2pos_el-TP2
FP2 <- S2neg_el-TN2
S3sens_el <- 0.711
S3spec_el <- 1
S3pos_el <- 45
S3neg_el <- 22
TP3 <- round(S3sens_el*S3pos_el)
TN3 <- round(S3spec_el*S3neg_el)
FN3 <- S3pos_el-TP3
FP3 <- S3neg_el-TN3
S4sens_el <- 0.600
S4spec_el <- 0.99
```

```
TP4 <- 55
TN4 <- 294
FN4 <- 37
FP4 <- 2

AuditC5_el <- data.frame(TP = c(TP1, TP2, TP3, TP4),
                FN = c(FN1, FN2, FN3, FN4),
                FP = c(FP1, FP2, FP3, FP4),
                TN = c(TN1, TN2, TN3, TN4))
AuditC5_el$names <- c("Study 1", "Study2", "Study 3", "Study 4")

sens_logit_el <- metaprop(AuditC5_el$TP, AuditC5_el$TP + AuditC5_el$FN, comb.fixed=F, comb.random=T,
sm="PLOGIT", method = "GLMM", method.ci="CP", studlab=AuditC5_el$names)

spec_logit_el <- metaprop(AuditC5_el$TN, AuditC5_el$TN + AuditC5_el$FP, comb.fixed=FALSE,
comb.random=TRUE, sm="PLOGIT", method.ci="CP", studlab=AuditC5_el$names)
```

### Outputs

```
        proportion         95%-CI
Study 1      0.864 [0.651; 0.971]
Study2       0.900 [0.735; 0.979]
Study 3      0.711 [0.557; 0.836]
Study 4      0.598 [0.490; 0.699]

Number of studies combined: k = 4

                     proportion         95%-CI
Random effects model      0.769 [0.611; 0.876]

Quantifying heterogeneity:
 tau^2 = 0.3653; tau = 0.6044; I^2 = 74.4% [28.8%; 90.8%]; H = 1.98 [1.19; 3.30]

Test of heterogeneity:
     Q d.f. p-value             Test
 11.74    3  0.0083        Wald-type
 14.58    3  0.0022 Likelihood-Ratio
```

**Figure S4: Output of the metaprop function for the estimation of the sensitivity of the ELISA**

```
        proportion         95%-CI
Study 1      0.962 [0.870; 0.995]
Study2       1.000 [0.955; 1.000]
Study 3      1.000 [0.846; 1.000]
Study 4      0.993 [0.976; 0.999]

Number of studies combined: k = 4

                     proportion         95%-CI
Random effects model      0.991 [0.977; 0.997]

Quantifying heterogeneity:
 tau^2 = 0; tau = 0; I^2 = 0.0% [0.0%; 84.7%]; H = 1.00 [1.00; 2.56]

Test of heterogeneity:
    Q d.f. p-value             Test
 3.00    3  0.3917        Wald-type
 4.76    3  0.1904 Likelihood-Ratio
```

**Figure S5: Output of the metaprop function for the estimation of the specificity of the ELISA**

### Adjustment 2 – R Code

BLCM model definition:

```
model{

	## Observation layer:

	# Complete observations (N=825):
	for(p in 1:Populations){
		Tally_RR[1:4,p] ~ dmulti(prob_RR[1:4,p], N_RR[p])

		prob_RR[1:4,p] <- se_prob[1:4,p] + sp_prob[1:4,p]
	}

	## Observation probabilities:

	for(p in 1:Populations){

		# Probability of observing RDT- ELISA- from a true positive::
		se_prob[1,p] <- prev[p] * ((1-se[1])*(1-se[2]) +covse12)
		# Probability of observing RDT- ELISA- from a true negative::
		sp_prob[1,p] <- (1-prev[p]) * (sp[1]*sp[2] +covsp12)

		# Probability of observing RDT+ ELISA- from a true positive::
		se_prob[2,p] <- prev[p] * (se[1]*(1-se[2]) -covse12)
		# Probability of observing RDT+ ELISA- from a true negative::
		sp_prob[2,p] <- (1-prev[p]) * ((1-sp[1])*sp[2] -covsp12)

		# Probability of observing RDT- ELISA+ from a true positive::
		se_prob[3,p] <- prev[p] * ((1-se[1])*se[2] -covse12)
		# Probability of observing RDT- ELISA+ from a true negative::
		sp_prob[3,p] <- (1-prev[p]) * (sp[1]*(1-sp[2]) -covsp12)

		# Probability of observing RDT+ ELISA+ from a true positive::
		se_prob[4,p] <- prev[p] * (se[1]*se[2] +covse12)
		# Probability of observing RDT+ ELISA+ from a true negative::
		sp_prob[4,p] <- (1-prev[p]) * ((1-sp[1])*(1-sp[2]) +covsp12)
	}

	## Priors:
	# Prevalence in population [0,5):
	prev[1] ~ dbeta(1,1)
	# Prevalence in population [5,20):
	prev[2] ~ dbeta(1,1)
	# Prevalence in population [20,35):
	prev[3] ~ dbeta(1,1)
	# Prevalence in population [35,50):
	prev[4] ~ dbeta(1,1)
	# Prevalence in population [50,Inf):
	prev[5] ~ dbeta(1,1)

	# Sensitivity of RDT test:
	se[1] ~ dbeta(61.22,19.40)T(1-sp[1], )
	# Specificity of RDT test:
	sp[1] ~ dbeta(965.86, 10.74)

	# Sensitivity of ELISA test:
	se[2] ~ dbeta(21.77, 7.24)T(1-sp[2], )
	# Specificity of ELISA test:
```

```
            sp[2] ~ dbeta(332.21, 4.01)


            # Covariance in sensitivity between RDT and ELISA tests:
            # covse12 ~ dunif( (se[1]-1)*(1-se[2]) , min(se[1],se[2]) - se[1]*se[2] )  ## if the sensitivity of these tests may be correlated
             covse12 <- 0  ## if the sensitivity of these tests can be assumed to be independent
            # Covariance in specificity between RDT and ELISA tests:
            # covsp12 ~ dunif( (sp[1]-1)*(1-sp[2]) , min(sp[1],sp[2]) - sp[1]*sp[2] )  ## if the specificity of these tests may be correlated
             covsp12 <- 0  ## if the specificity of these tests can be assumed to be independent

}

#monitor# se, sp, prev, covse12, covsp12

## Inits:
inits{
"se" <- c(0.5, 0.99)
"sp" <- c(0.99, 0.75)
"prev" <- c(0.05, 0.95, 0.05, 0.95, 0.05)
# "covse12" <- 0
# "covsp12" <- 0
}
inits{
"se" <- c(0.99, 0.5)
"sp" <- c(0.75, 0.99)
"prev" <- c(0.95, 0.05, 0.95, 0.05, 0.95)
# "covse12" <- 0
# "covsp12" <- 0
}

## Data:
data{
"Populations" <- 5
"N_RR" <- c(60, 312, 214, 136, 103)
"Tally_RR" <- structure(c(44, 1, 9, 6, 161, 25, 56, 70, 126, 16, 36, 36, 68, 10, 24, 34, 44, 8, 13, 38), .Dim = c(4, 5))
}


##Model call

# Set different starting values for the three different chains to assess convergence
inits1 = list(".RNG.name" ="base::Mersenne-Twister",
            ".RNG.seed" = 100022)
inits2 = list(".RNG.name" ="base::Mersenne-Twister",
            ".RNG.seed" = 300022)
inits3 = list(".RNG.name" ="base::Mersenne-Twister",
            ".RNG.seed" = 500022)

run.jags("./autohw.bug",
        inits=list(inits1, inits2, inits3),
        n.chains = 3,
        burnin = 10000,
        sample = 100000,
        adapt = 1000,
       )

## Calculating the adjusted seroprevalence
```

```
adj_bay_serop_overall <- epi.prev(nrow(sero[test_result_cat=="Positive", ]), method = "c-p",
                    nrow(sero[!is.na(test_result_cat) & outcome == "done", ]),
                    se=0.619, sp=0.989)

adj_bay_serop_o50 <- epi.prev(nrow(sero[age_group == "[50,Inf)" & test_result_cat=="Positive", ]),
                    nrow(sero[age_group == "[50,Inf)" & !is.na(test_result_cat) & outcome == "done", ]),
                    0.619, 0.989)
adj_bay_serop_35_50 <- epi.prev(nrow(sero[age_group == "[35,50)" & test_result_cat=="Positive", ]),
                    nrow(sero[age_group == "[35,50)" & !is.na(test_result_cat) & outcome == "done", ]),
                    0.619, 0.989)
adj_bay_serop_20_35 <- epi.prev(nrow(sero[age_group == "[20,35)" & test_result_cat=="Positive", ]),
                    nrow(sero[age_group == "[20,35)" & !is.na(test_result_cat) & outcome == "done", ]),
                    0.619, 0.989)
adj_bay_serop_5_20 <- epi.prev(nrow(sero[age_group == "[5,20)" & test_result_cat=="Positive", ]),
                    nrow(sero[age_group == "[5,20)" & !is.na(test_result_cat) & outcome == "done", ]),
                    0.619, 0.989)
adj_bay_serop_0_5 <- epi.prev(nrow(sero[age_group == "[0,5)" & test_result_cat=="Positive", ]),
                    nrow(sero[age_group == "[0,5)" & !is.na(test_result_cat) & outcome == "done", ]),
                    0.619, 0.989)
```

### BLCM Output

```
JAGS model summary statistics from 300000 samples (chains = 3; adapt+burnin = 11000):

       Lower95  Median   Upper95   Mean     SD Mode     MCerr        MC%ofSD SSeff
se[1]  0.5682   0.61911  0.66909   0.61905  0.025775  -- 0.00015081   0.6     29209
sp[1]  0.98247  0.9894   0.99494   0.98908  0.0032768 -- 0.000018919  0.6     30000
se[2]  0.72266  0.77446  0.82388   0.77415  0.025912  -- 0.0001496    0.6     30000
sp[2]  0.9697   0.98578  0.99721   0.98457  0.0075763 -- 0.000043742  0.6     30000

       AC.10        psrf
se[1]  0.0020527    0.99999
sp[1]  -0.000081098 1.0001
se[2]  0.00016124   1.0002
sp[2]  0.00051855   1.0001
```

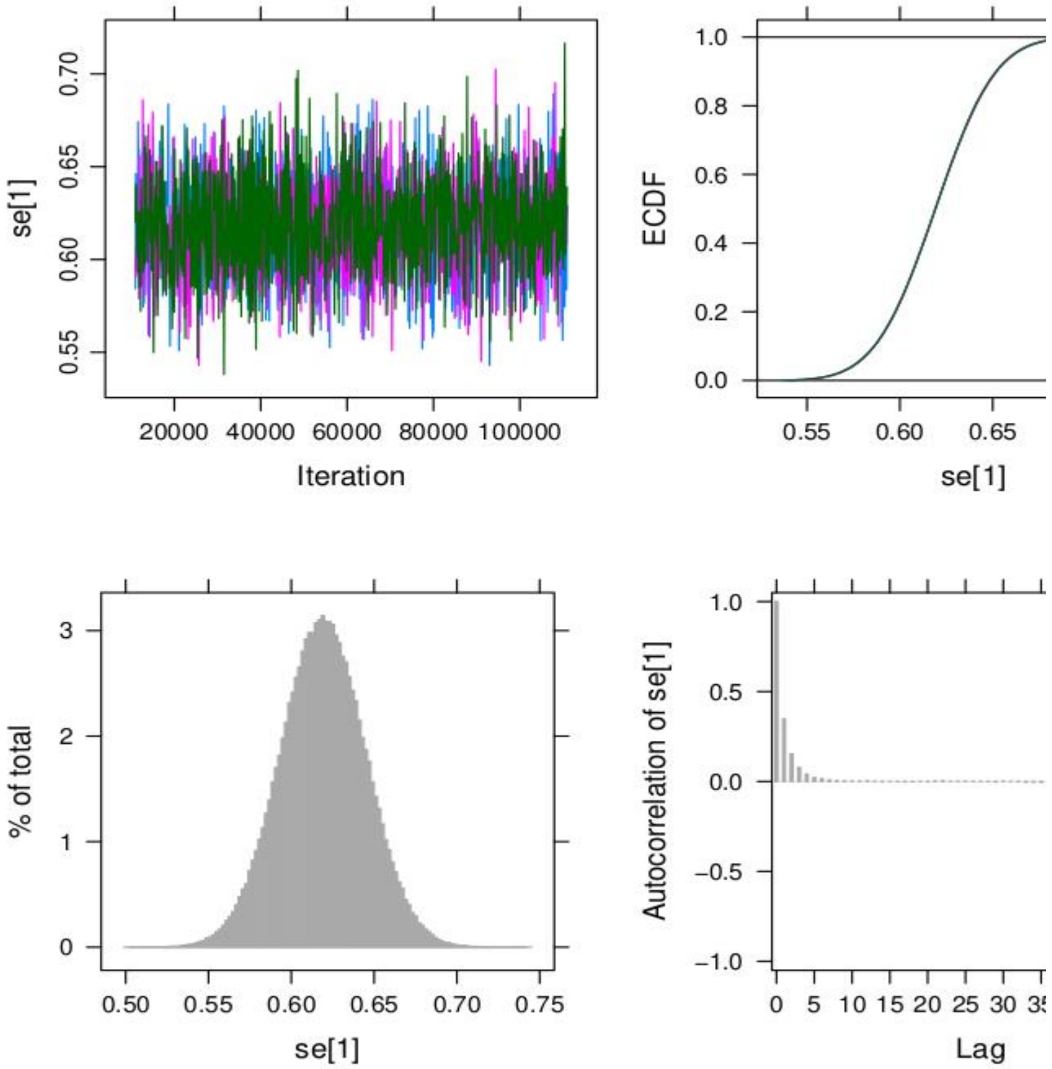


**Figure S6: Output of the runjags function to estimate the sensitivity of the RST. Upper left: Trace plot of the 3 chains showing to be stationary; Upper right: ECDF plot of the three chains overlapping; Bottom left: Histograms of the combined chains; Bottom right: Autocorrelation plot**

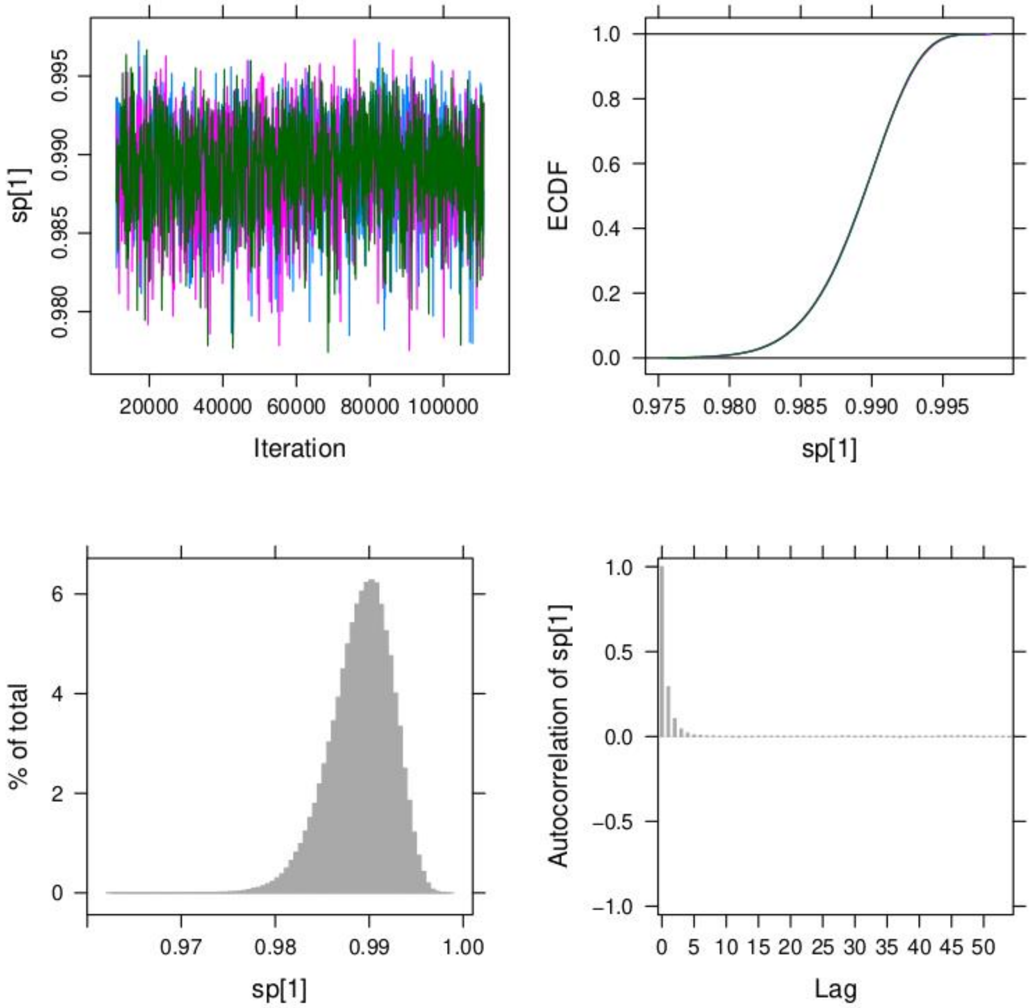


Figure

**Figure S7: Output of the runjags function to estimate the specificity of the RST. Upper left: Trace plot of the 3 chains showing to be stationary; Upper right: ECDF plot of the three chains overlapping; Bottom left: Histograms of the combined chains; Bottom right: Autocorrelation plot**

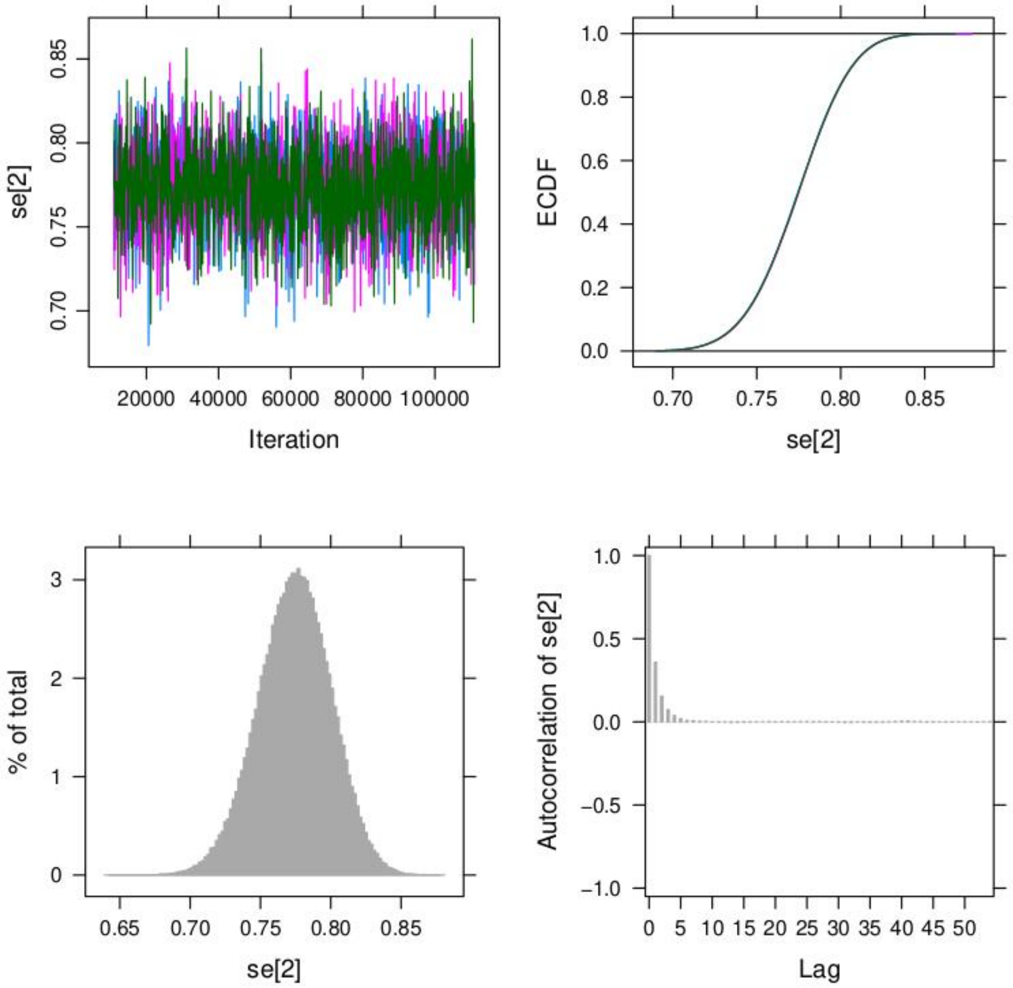


**Figure S8: Output of the runjags function to estimate the sensitivity of the ELISA. Upper left: Trace plot of the 3 chains showing to be stationary; Upper right: ECDF plot of the three chains overlapping; Bottom left: Histograms of the combined chains; Bottom right: Autocorrelation plot**

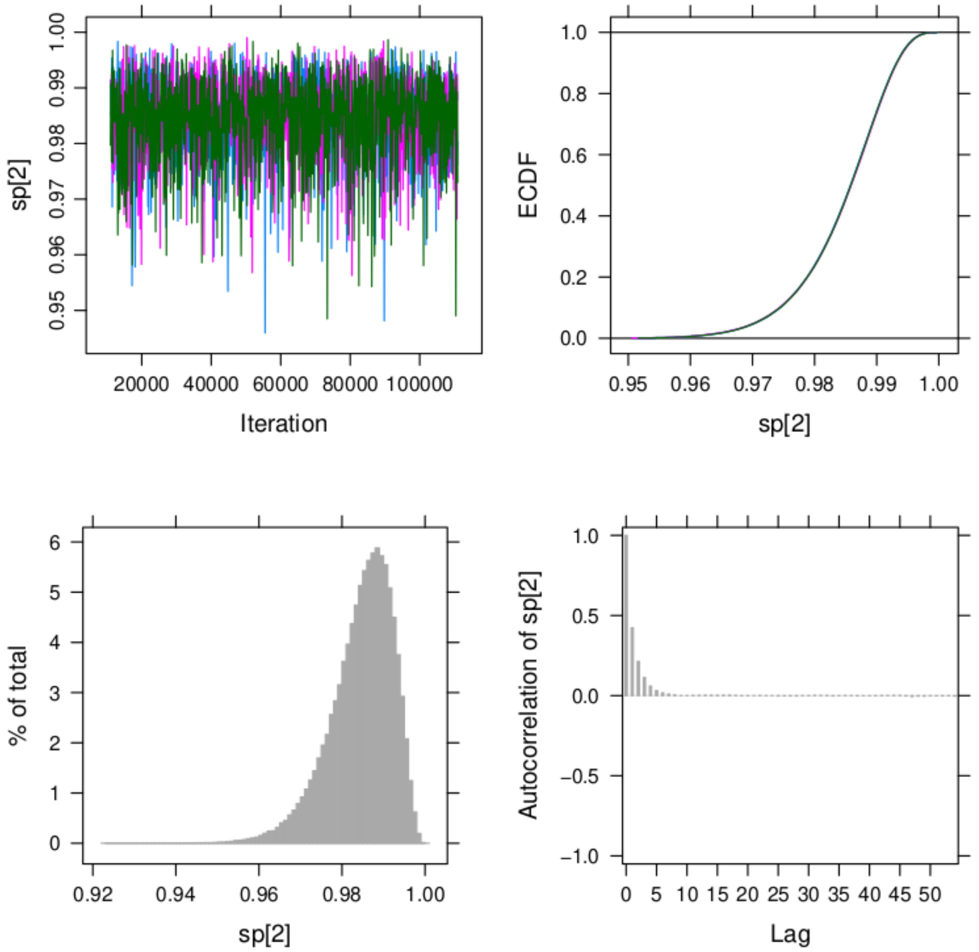


**Figure S9: Output of the runjags function to estimate the specificity of the ELISA. Upper left: Trace plot of the 3 chains showing to be stationary; Upper right: ECDF plot of the three chains overlapping; Bottom left: Histograms of the combined chains; Bottom right: Autocorrelation plot**

| Causes of death/Age group | <20 years | 20-34 years | 35-49 years | ≥50 years | Total |
|---|---|---|---|---|---|
| **Accident / Trauma** | 12.9% (4) | 22.6% (7) | 19.4% (6) | 45.2% (14) | 100.0% (31) |
| **Cancer** | 4.3% (1) | 8.7% (2) | 21.7% (5) | 65.2% (15) | 100.0% (23) |
| **Chronic kidney disease** | 0.0% (0) | 0.0% (0) | 0.0% (0) | 100.0.0% (1) | 100.0.0% (1) |
| **COVID-19** | 11.1% (1) | 0.0% (0) | 11.1% (1) | 77.8% (7) | 100.0% (9) |
| **Diarrhoea** | 14.3% (1) | 14.3% (1) | 0.0% (0) | 71.4% (5) | 100.0% (7) |
| **Don't know** | 13.0% (7) | 5.6% (3) | 7.4% (4) | 74.1% (40) | 100.0% (54) |
| **Isolated fever / Malaria** | 7.7% (1) | 15.4% (2) | 15.4% (2) | 61.5% (8) | 100.0% (13) |
| **Malnutrition** | 50.0% (1) | 0.0% (0) | 0.0% (0) | 50.0% (1) | 100.0% (2) |
| **NCD** | 1.2% (1) | 2.3% (2) | 14.0% (12) | 82.6% (71) | 100.0% (86) |
| **Neonatal death/disease** | 91.7% (11) | 8.3% (1) | 0.0% (0) | 0.0% (0) | 100.0% (12) |
| **Others** | 11.5% (6) | 11.5% (6) | 13.5% (7) | 63.5% (33) | 100.0% (52) |
| **Pregnancy/childbirth related deaths** | 50.0% (3) | 33.3% (2) | 16.7% (1) | 0.0% (0) | 100.0% (6) |
| **Respiratory diseases** | 29.4% (5) | 5.9% (1) | 11.8% (2) | 52.9% (9) | 100.0% (17) |
| **Unknown** | 0.0% (0) | 0.0% (0) | 0.0% (0) | 100.0% (1) | 100.0% (1) |
| **Violence** | 20% (1) | 60.0% (3) | 0.0% (0) | 20.0% (1) | 100.0% (5) |
| **Total** | 13.5% (43) | 9.4% (30) | 12.5% (40) | 64.6% (206) | 100.0% (319) |

**Table S1. Summary of causes of deaths among the different age groups**

NCD=Non-communicable disease.

| *Causes of death [% (n)]* | *Age group* | | | | | | | | | |
|---|---|---|---|---|---|---|---|---|---|---|
| | **<20 years** | | **20-34 years** | | **35-49 years** | | **≥50 years** | | **Total** | |
| | **PREP** | **PAND** | **PREP** | **PAND** | **PREP** | **PAND** | **PREP** | **PAND** | **PREP** | **PAND** |
| ***Accident / Trauma*** | 5% (1) | 13% (3) | 10% (1) | 30% (6) | 31% (5) | 4% (1) | 8% (6) | 6% (8) | 10% (13) | 9% (18) |
| ***Cancer*** | 5% (1) | 0% (0) | 10% (1) | 5% (1) | 12% (2) | 12% (3) | 8% (6) | 7% (9) | 8% (10) | 7% (13) |
| ***Chronic kidney disease*** | 0% (0) | 0% (0) | 0% (0) | 0% (0) | 0% (0) | 0% (0) | 1% (1) | 0% (0) | 1% (1) | 0% (0) |
| ***COVID-19*** | 0% (0) | 4% (1) | 0% (0) | 0% (0) | 0% (0) | 4% (1) | 0% (0) | 6% (7) | 0% (0) | 5% (9) |
| ***Diarrhoea*** | 0% (0) | 4% (1) | 10% (1) | 0% (0) | 0% (0) | 0% (0) | 1% (1) | 3% (4) | 2% (2) | 3% (5) |
| ***Don't know*** | 10% (2) | 22% (5) | 10% (1) | 10% (2) | 12% (2) | 8% (2) | 24% (19) | 17% (21) | 19% (24) | 16% (30) |
| ***Isolated fever / Malaria*** | 5% (1) | 0% (0) | 10% (1) | 5% (1) | 6% (1) | 4% (1) | 2% (2) | 5% (6) | 4% (5) | 4% (8) |
| ***Malnutrition*** | 5% (1) | 0% (0) | 0% (0) | 0% (0) | 0% (0) | 0% (0) | 0% (0) | 1% (1) | 1% (1) | 1% (1) |
| ***NCD*** | 0% (0) | 4% (1) | 0% (0) | 10% (2) | 31% (5) | 29% (7) | 35% (28) | 34% (43) | 26% (33) | 27% (53) |
| ***Neonatal death/disease*** | 30% (6) | 22% (5) | 0% (0) | 5% (1) | 0% (0) | 0% (0) | 0% (0) | 0% (0) | 5% (6) | 3% (6) |
| ***Others*** | 10% (2) | 17% (4) | 20% (2) | 20% (4) | 6% (1) | 25% (6) | 19% (15) | 14% (18) | 16% (20) | 17% (32) |
| ***Pregnancy/childbirth related*** | 10% (2) | 4% (1) | 20% (2) | 0% (0) | 0% (0) | 4% (1) | 0% (0) | 0% (0) | 3% (4) | 1% (2) |
| ***Respiratory infections*** | 20% (4) | 4% (1) | 0% (0) | 5% (1) | 0% (0) | 8% (2) | 2% (2) | 6% (7) | 5% (6) | 6% (11) |
| ***Unknown*** | 0% (0) | 0% (0) | 0% (0) | 0% (0) | 0% (0) | 0% (0) | 0% (0) | 1% (1) | 0% (0) | 1% (1) |
| ***Violence*** | 0% (0) | 4% (1) | 10% (1) | 10% (2) | 0% (0) | 0% (0) | 0% (0) | 1% (1) | 1% (1) | 2% (4) |
| ***Total (n)*** | 100% (20) | 100% (23) | 100% (10) | 100% (20) | 100% (16) | 100% (24) | 100% (80) | 100% (126) | 100% (126) | 100% (193) |

**Table S2. Summary of cause of deaths by period and age group**

PREP=Pre-pandemic period from January 1, 2019, until February 29, 2020. PAND=Pandemic period from March 1, 2020, until the end of the survey. NCD=Non-communicable disease.

| Symptom | Pre-pandemic % (n) | Pandemic % (n) | P-value |
|---|---|---|---|
| **Fever** | 12.7% (16) | 16.6% (32) | <0.001 |
| **Cough** | 5.6% (7) | 9.3% (18) | 0.288 |
| **Shortness of breath** | 15.1% (19) | 14.5% (28) | 0.873 |
| **Weakness/Fatigue** | 11.1% (14) | 15.5% (30) | 0.32 |
| **Extreme fatigue** | 4.8% (6) | 11.4% (22) | 0.044 |
| **Headaches** | 6.3% (8) | 10.4% (20) | 0.233 |
| **Muscle pain** | 4.8% (6) | 11.9% (23) | 0.044 |
| **Sore throat** | 2.4% (3) | 1.6% (3) | 0.684 |
| **Runny nose** | 0.8% (1) | 0.5% (1) | >0.999 |
| **Loss appetite** | 4.8% (6) | 10.4% (20) | 0.094 |
| **Diarrhoea** | 5.6% (7) | 5.7% (11) | >0.999 |
| **Change mental state** | 4.0% (5) | 2.6% (5) | 0.524 |
| **Loss taste/odour** | 0.8% (1) | 0.5% (1) | >0.999 |
| **No symptoms** | 40.5% (51) | 34.7% (67) | 0.343 |
| **Don't know** | 19.8% (25) | 20.7% (40) | 0.888 |

**Table S3. Summary of symptoms reported prior to deaths in pre-pandemic and pandemic periods**

| **Location** | **Pre-pandemic % (n)** | **Pandemic % (n)** | **P-value** |
|---|---|---|---|
| **Asthma** | 0.0% (0) | 3.6% (3) | 0.553 |
| **Autoimmune disease (polyarthritis, Crohn's disease, lupus, multiple sclerosis…)** | 2.1% (1) | 2.4% (2) | >0.999 |
| **Cancer** | 6.2% (3) | 7.1% (6) | >0.999 |
| **Chronic lung disease** | 0.0% (0) | 1.2% (1) | >0.999 |
| **Congestive heart failure** | 6.2% (3) | 1.2% (1) | 0.136 |
| **Coronary heart disease** | 6.2% (3) | 8.3% (7) | 0.747 |
| **Current smoker** | 0.0% (0) | 1.2% (1) | >0.999 |
| **Diabetes** | 22.9% (11) | 22.6% (19) | >0.999 |
| **Hepatitis B** | 0.0% (0) | 1.2% (1) | >0.999 |
| **Hypertension** | 33.3% (16) | 28.6% (24) | 0.563 |
| **Kidney Disease** | 10.4% (5) | 8.3% (7) | 0.757 |
| **Other** | 12.5% (6) | 14.3% (12) | >0.999 |

**Table S4. Reported comorbidities for deaths during the pre-pandemic and pandemic periods**

| | Pre-pandemic<br>% (n) | Pandemic<br>% (n) | P-value |
|---|---|---|---|
| **Access to health care** | | | |
| Health Centre | 5.0% (4) | 8.3% (10) | 0.413 |
| Hospital | 82.5% (66) | 75.0% (90) | 0.227 |
| Other | 1.2% (1) | 0.8% (1) | >0.999 |
| Pharmacist | 0.0% (0) | 0.8% (1) | >0.999 |
| Self-medication: modern medicine | 10.0% (8) | 15.0% (18) | 0.392 |
| Self-medication: traditional medicine | 1.2% (1) | 0.0% (0) | 0.400 |
| **Location of death** | | | |
| Don't know | 0.8% (1) | 0.0% (0) | 0.395 |
| Health Centre | 0.8% (1) | 1.6% (3) | >0.999 |
| Home | 43.7% (55) | 47.2% (91) | 0.567 |
| Hospital | 50% (63) | 45.1% (87) | 0.423 |
| On the way to the health centre/hospital | 0.8% (1) | 3.1% (6) | 0.251 |
| Other | 4% (5) | 3.1% (6) | 0.758 |

**Table S5. Summary of access to health care type and location of death for the pre-pandemic and pandemic periods**

| Age group | Consented % (n) | | Refused % (n) | | Absent % (n) | |
|---|---|---|---|---|---|---|
| | female | male | female | male | female | male |
| **<5 years** | 43.5% (130) | 56.5% (169) | 58.4% (66) | 41.6% (47) | 48.3% (14) | 51.7% (15) |
| **5-19 years** | 54.0% (424) | 46.0% (361) | 46.4% (123) | 53.6% (142) | 43.5% (124) | 56.5% (161) |
| **20-34 years** | 60.9% (383) | 39.1% (246) | 40.9% (74) | 59.1% (107) | 26.5% (59) | 73.5% (164) |
| **35-49 years** | 63.5% (217) | 36.5% (125) | 26.0% (20) | 74.0% (57) | 27.0% (27) | 73.0% (73) |
| **≥50 years** | 49.8% (159) | 50.2% (160) | 44.6% (37) | 55.4% (46) | 24.1% (19) | 75.9% (60) |
| **Total** | **62.3% (2374)** | | **18.9% (719)** | | **18.8% (716)** | |

**Table S6. Status of acceptance to participate in serology survey by age group and gender**

| | Crude OR (95% CI) | Adjusted OR | |
|---|---|---|---|
| | | OR (95% CI) | P-value |
| **Age*** | 1.01 (1.01-1.02) | 1.01 (1.00-1.02) | 0.001 |
| **Sex**** | 0.82 (0.66-1.02) | 0.84 (0.67-1.05) | 0.127 |
| **Past medical history***** | 1.30 (1.00-1.69) | 1.06 (0.79-1.42) | 0.703 |
| **Not exposed to case in household****** | 0.60 (0.48-0.74) | 0.62 (0.50-0.77) | < 0.001 |

**Table S7. The risk factor in association with a positive SARS-Cov-2 rapid antibody test**
*Continuous variable. **Reference value is female; ***Reference value has a medical history. ****Reference value has not been exposed to a case within the household. OR=odds ratio.

| Age group | Positive IgG<br>% (n) | Positive IgG and IgM<br>% (n) | Positive IgM<br>% (n) | Total<br>% (n) |
|---|---|---|---|---|
| **<5 years** | 92.9% (52) | 1.8% (1) | 5.4% (3) | 100.% (56) |
| **5–19 years** | 90.8% (218) | 7.9% (19) | 1.2% (3) | 100.% (240) |
| **20–34 years** | 78.5% (175) | 12.1% (27) | 9.4% (21) | 100.% (223) |
| **35–49 years** | 83.0% (112) | 12.6% (17) | 4.4% (6) | 100.% (135) |
| **≥50 years** | 83.8% (134) | 13.8% (22) | 2.5% (4) | 100.% (160) |
| **Total** | 84.9% (691) | 10.6% (86) | 4.5% (37) | 100.% (814) |

**Table S8. Summary of positive rapid serologic testing by the antibody status**

| | ELISA negative<br>% (n) | ELISA positive<br>% (n) | Total<br>% (n) |
|---|---|---|---|
| **RST negative** | 76.2% (443) | 23.8% (138) | 100.0% (581) |
| **RST positive** | 24.6% (60) | 75.4% (184) | 100.0% (244) |
| **Total** | 61.0% (503) | 39.0% (322) | 100.0% (825) |

**Table S9. Summary of ELISA and RST results**

ELISA=enzyme-linked immunosorbent assay. RST=rapid serologic test.

| Age group | Female<br>% (95% CI) | Male<br>% (95% CI) | Total<br>% (95% CI) |
|---|---|---|---|
| **<5 years** | 22.9% (17.0–30.1) | 15.2% (9.7–23.1) | 18.6% (14.0–24.2) |
| **5–19 years** | 32.1% (27.0–37.6) | 28.7% (23.3–34.8) | 30.5% (26.3–35.1) |
| **20–34 years** | 32.0% (27.3–37.1) | 40.6% (33.0–48.7) | 35.3% (30.7–40.3) |
| **35–49 years** | 40.5% (34.5–46.9) | 37.4% (28.6–47.1) | 39.4% (34.9–44.1) |
| **≥50 years** | 50.4% (41.5–59.2) | 50.1% (42.8–57.4) | 50.2% (44.0–56.5) |

**Table S10. Prevalence of SARS-CoV-2 antibodies based on rapid serologic test by age group and sex**
CI=confidence interval.

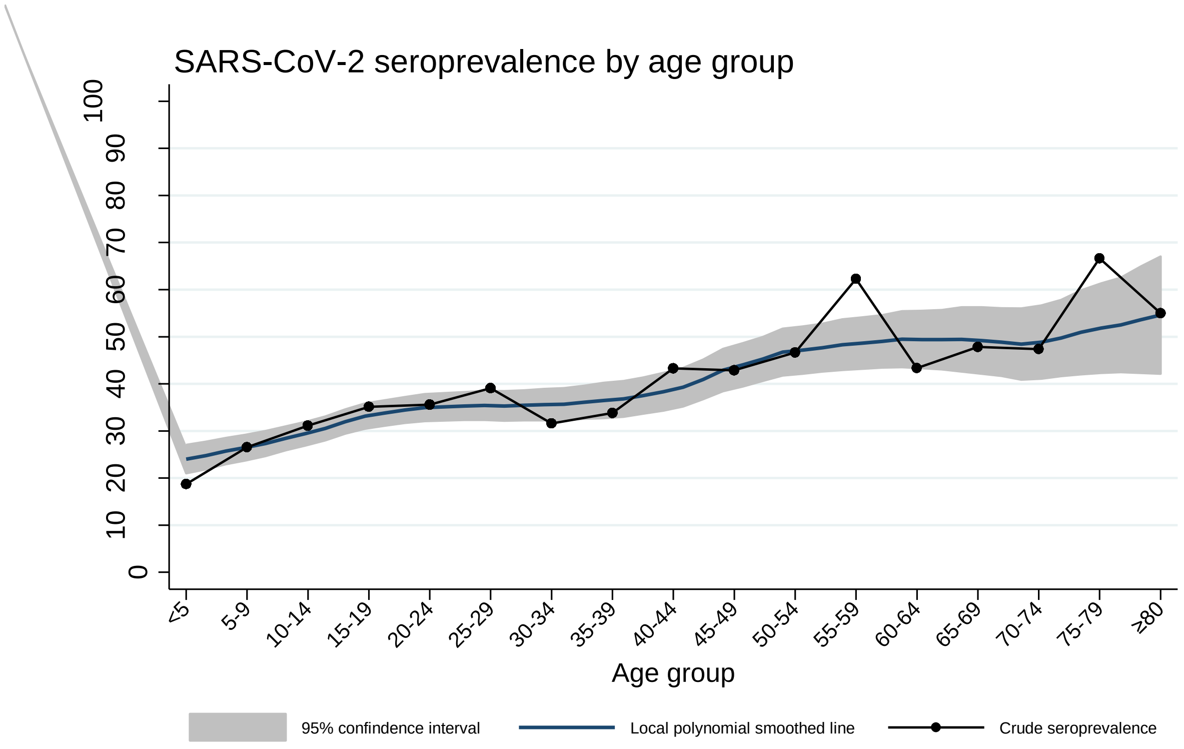


**Figure S10. SARS-CoV-2 seroprevalence by age group**
